# Bias correction methods for test-negative designs in the presence of misclassification

**DOI:** 10.1101/19002691

**Authors:** Akira Endo, Sebastian Funk, Adam J. Kucharski

## Abstract

The test-negative design has become a standard approach for vaccine effectiveness studies. However, previous studies suggested that it may be more sensitive than other designs to misclassification of disease outcome caused by imperfect diagnostic tests. This could be a particular limitation in vaccine effectiveness studies where simple tests (e.g. rapid influenza diagnostic tests) are used for logistical convenience. To address this issue, we derived a mathematical representation of the test-negative design with imperfect tests, then developed a bias correction framework for possible misclassification. Test-negative design studies usually include multiple covariates other than vaccine history to adjust potential confounders; our methods can also address multivariate analyses and be easily coupled with existing estimation tools. We validated the performance of these methods using simulations of common scenarios for vaccine efficacy and were able to obtain unbiased estimates in a variety of parameter settings.

## 1 Introduction

Vaccine effectiveness (VE) is typically estimated as the vaccine-induced risk reduction of the target disease (TD) and has been traditionally studied by the cohort or case-control designs. However, the test-negative design (TND) is becoming a popular alternative design for vaccine effectiveness (VE) studies [1, 2]. This is a modified version of the case-control study with an alternative definition of the control group; traditional case-control studies usually define controls as non-disease individuals in the study population, while the TND studies use individuals with similar symptoms to the target disease but presenting negative test results (i.e., patients of non-target diseases; ND). The test-negative design can therefore minimise ascertainment bias by including only medically-attended patients in both case and control groups. Many TND studies have focused on influenza vaccination, but recent studies have also targeted other diseases including pneumococcal disease [3, 4] and rotavirus disease [5, 6, 7].

Despite its increasing popularity, TND can be more sensitive than other study designs to misclassification of disease outcome. Multiple studies have shown that VE is underestimated when the diagnostic tests used in the study are imperfect (i.e. have a sensitivity and/or a specificity less than 100%) [8, 9, 10]. This can be a particular issue when simple tests (e.g. rapid diagnostic tests) are used for logistical convenience, as simple tests tend to have lower diagnostic performance than more advanced tests (e.g. polymerase chain reaction; PCR). Previous studies evaluated the expected degree of bias and concluded that specificity had a more important effect on bias than sensitivity [8, 9, 10]. These findings appear to support the use of rapid tests, despite limited sensitivity, because the specificity of these tests is typically high [2]. However, theoretical studies to date have been based on specific assumptions about efficacy and pathogen epidemiology; it is therefore unclear whether such conclusions hold for all plausible combinations of scenarios.

If a study is expected to generate a non-negligible bias in estimation, such bias needs to be assessed and—if possible— corrected before the estimate is reported. Greenland [11] proposed a bias correction method for cohort studies where the sensitivity and specificity of the test are known (or at least assumed). However, this method cannot apply to case-control studies because of differential recruitment, whereby the probability of recruiting (test-positive) cases and (negative) controls may be different. Although TND studies are often considered to be special cases of case-control studies, they are free from the issue of differential recruitment because the recruitment and classification are mutually-independent. This means that, while Greenland’s method does not apply to TND, another type of bias correction may still be possible. For example, De Smedt et al. have characterised the misclassification bias in VE in the test-negative design in a simulation study [3]. One limitation of this approach is it relies on the unobserved “true” disease risk being known, where in reality this is not usually measurable in field studies. As a result, bias correction methods for TND studies that are directly applicable to field data have not yet been proposed. Moreover, previous analysis of misclassification bias has not considered the impact of multivariate analysis, where potential confounders (e.g. age and sex) are also included in the model used to estimate VE.

To address these issues, we develop a bias correction method for the test-negative VE studies that uses only data commonly available in field studies. We also apply these methods to multivariate analyses. As our approach uses the so-called multiple overimputation framework (generalisation of multiple imputation) [12], it can easily be coupled with a wide range of estimation tools without modifying their inside algorithms. Finally, we evaluate the performance of our methods by simulations of plausible epidemiological scenarios.

## 2 Methods and results

### 2.1 Characterising bias in test negative design studies

First, we consider the case where only vaccination history is included as a risk factor of acquiring the TD (i.e. the univariate setting). In this case, the (true) case counts can in theory be summarised in a two-by-two table as shown below:

|  | Vaccinated | Unvaccinated |
| --- | --- | --- |
| Target disease | $x_V$ | $x_U$ |
| Non-target disease | $y_V$ | $y_U$ |
| Subtotal | $S_V$ | $S_U$ |

Following the approach of Haber et al. (2015) [13], we consider four steps in the case reporting process: vaccination, onset of symptoms, seeking of medical care, and diagnosis. For simplicity, let us assume that occurrence of TD and ND are mutually independent, where their prevalences are represented as *r*_1_ and *r*_0_, respectively^1^. Let *v* be the vaccination coverage; in observational studies, vaccinated and unvaccinated population can have different likelihoods of seeking medical treatment (we denote such probabilities as *m*_*V*_ and *m*_*U*_, respectively). As our focus in the present study is the bias in VE estimation caused by imperfect tests, we made two key assumptions following Haber et al [13]: vaccination does not affect the risk of ND or the relative probability *µ* of medical attendance between TD and ND (which may reflect different disease severity between TD and ND). Namely, the study was assumed to be able to provide an unbiased VE estimate if tests are perfect. Based on these assumptions, we can classify the expected incidence in population *N* into four categories:

|  | Vaccinated | Unvaccinated |
| --- | --- | --- |
| Target disease | $v\gamma\mu m_V r_1 N$ | $(1-v)\mu m_U r_1 N$ |
| Non-target disease | $v m_V r_0 N$ | $(1-v)m_U r_0 N$ |
| Subtotal | $v m_V (\gamma\mu r_1 + r_0) N$ | $(1-v)m_U (\mu r_1 + r_0) N$ |

<sup>1</sup>It has been suggested that a possible violation of this assumption occur as a result of virus interference [14], but conclusive evidence for this is currently lacking [15, 16] and the effect on VE estimates may be limited in any case [17]

where *γ* is the relative risk of TD in the vaccinated population (i.e., *γ* = 1 − VE). *m*_*V*_ and *m*_*U*_ are the probabilities of the vaccinated and unvaccinated seeking medical care given ND (those given TD are *µm*_*V*_ and *µm*_*U*_, respectively). In TND studies, the (true) odds ratio corresponds to the relative risk *γ*.

Suppose that the true data in a TND study (*x*_*V*_, *y*_*V*_, *x*_*U*_, *y*_*U*_) is as described in the above tables. However, due to imperfect tests, we would instead expect to obtain the following observations:

|  | Vaccinated | Unvaccinated |
| --- | --- | --- |
| Test positive | $\alpha x_V + (1 - \beta)y_V$ | $\alpha x_U + (1 - \beta)y_U$ |
| Test negative | $(1 - \alpha)x_V + \beta y_V$ | $(1 - \alpha)x_U + \beta y_U$ |
| Subtotal | $S_V$ | $S_U$ |

where *α* and *β* are the sensitivity and specificity of the test, respectively. Denoting observed case counts with misclassification by *X* and *Y*, the process of diagnosis can be represented by the following matrix expression:

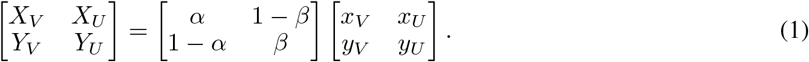

Matrix 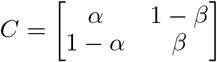 describes the conversion from the true disease state to the observed result. We hereafter refer to *C* as the classification matrix.

The observed odds ratio (subject to the misclassification bias) is therefore given as

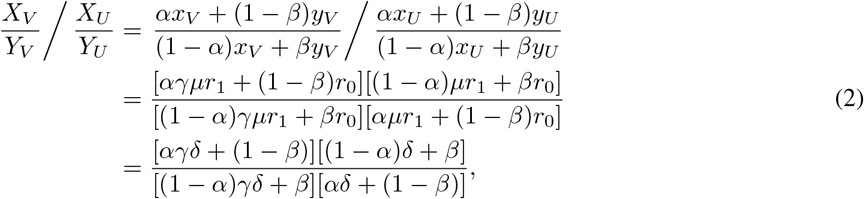

where 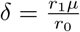 is the odds of the (medically-attended) target disease in the unvaccinated population.

We define bias in the VE estimate to be the absolute difference between the (raw) estimate and the true value. The expected bias *B* is a function of *α, β, γ* and *δ*:

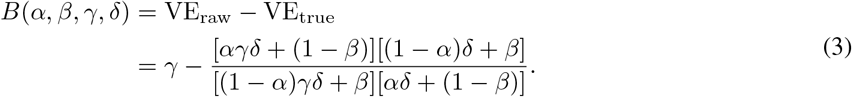

This suggests that the influence of sensitivity/specificity on the degree of bias varies depending on the case ratio *δ/*(1 + *δ*), i.e. the ratio between incidence of medical attendance for TD and ND in the study population (Figure 1). The degree of bias also depends on *γ* but is independent of *m*_*V*_ and *m*_*U*_. The degree of bias is largely determined by the test specificity when the case ratio is small, but the influence of sensitivity and specificity is almost equivalent with a case ratio of 0.6. It is notable that high specificity does not always assure that the bias is negligible. This may be true if specificity is strictly 100% and the case ratio is low to moderate, but a slight decline to 97% can cause a bias up to 10-15 percentage points. The effect of sensitivity is also non-negligible when the case ratio is high.

**Figure 1:**
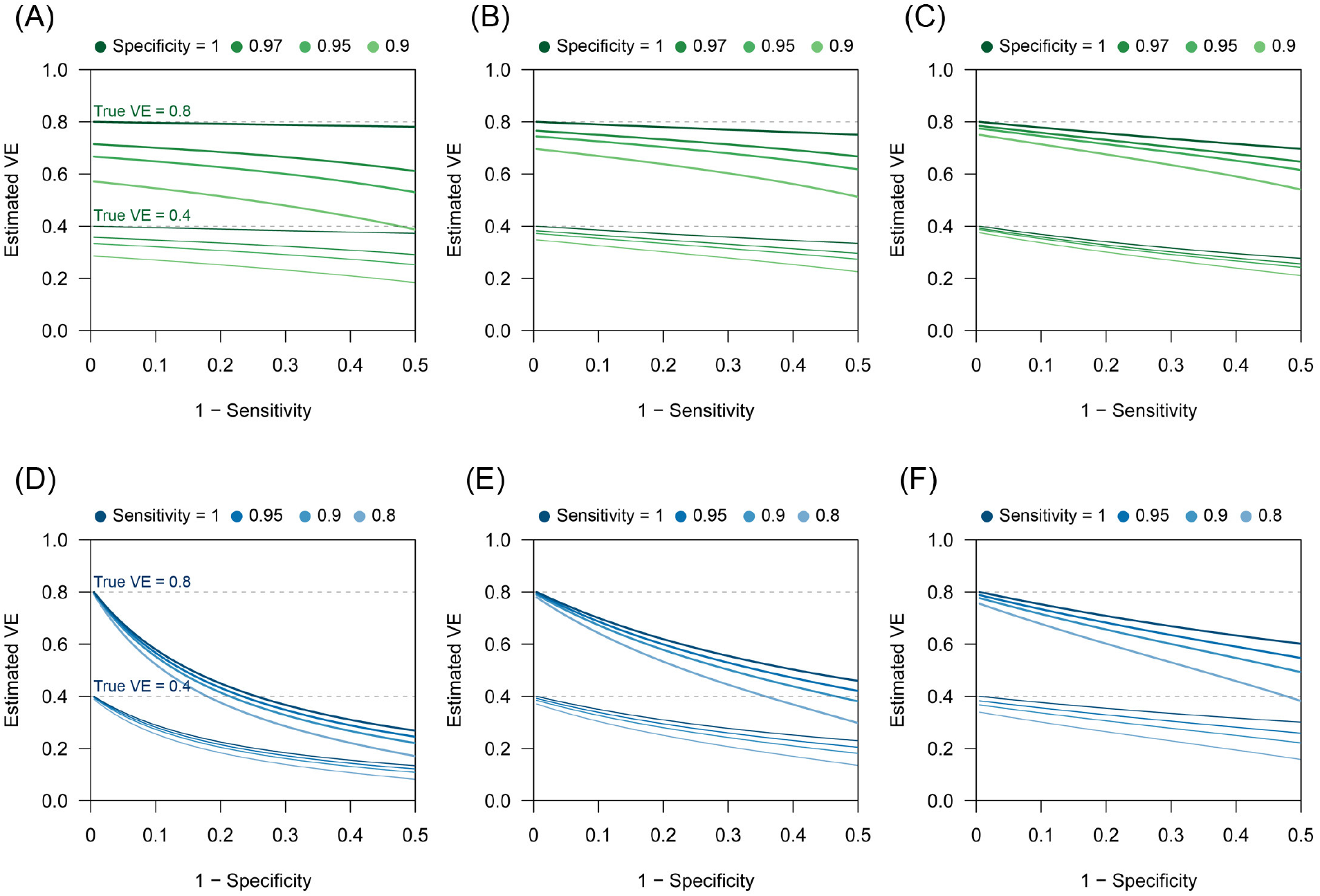
Bias in VE estimates caused by misclassification for different combinations of parameter values. (A) – (C) Estimated VE plotted against sensitivity. (A) True case ratio = 0.2 (B) 0.4 (C) 0.6. Each two sets of lines respectively correspond to different true VEs (80% and 40%, denoted by the dotted lines). (D) – (F) Estimated VE plotted against specificity. (D) True case ratio = 0.2 (E) 0.4 (F) 0.6.

When the expected bias is plotted against the case ratio with various combinations of test performance, we find that VE estimates can be substantially biased for certain case ratios (especially when the ratio is far from 1:1), even with reasonably high sensitivity and specificity (Figure 2A). In TND studies, researchers have no control over the case ratio because the study design requires that all tested individuals be included in the study. We found that the proportion of TD-positive patients in previous TND studies (retrieved from three systematic reviews [18, 19, 20]) varied considerably, ranging from 10% to 70% (Figure 2B) ^2^. Because of this large variation in the case ratio, it would be difficult to predict the degree of bias before data collection. Post-hoc assessment and correction therefore need to be considered. (Further analysis of the relationship between the degree of bias and parameter values can be found in the supplementary information.)

**Figure 2:**
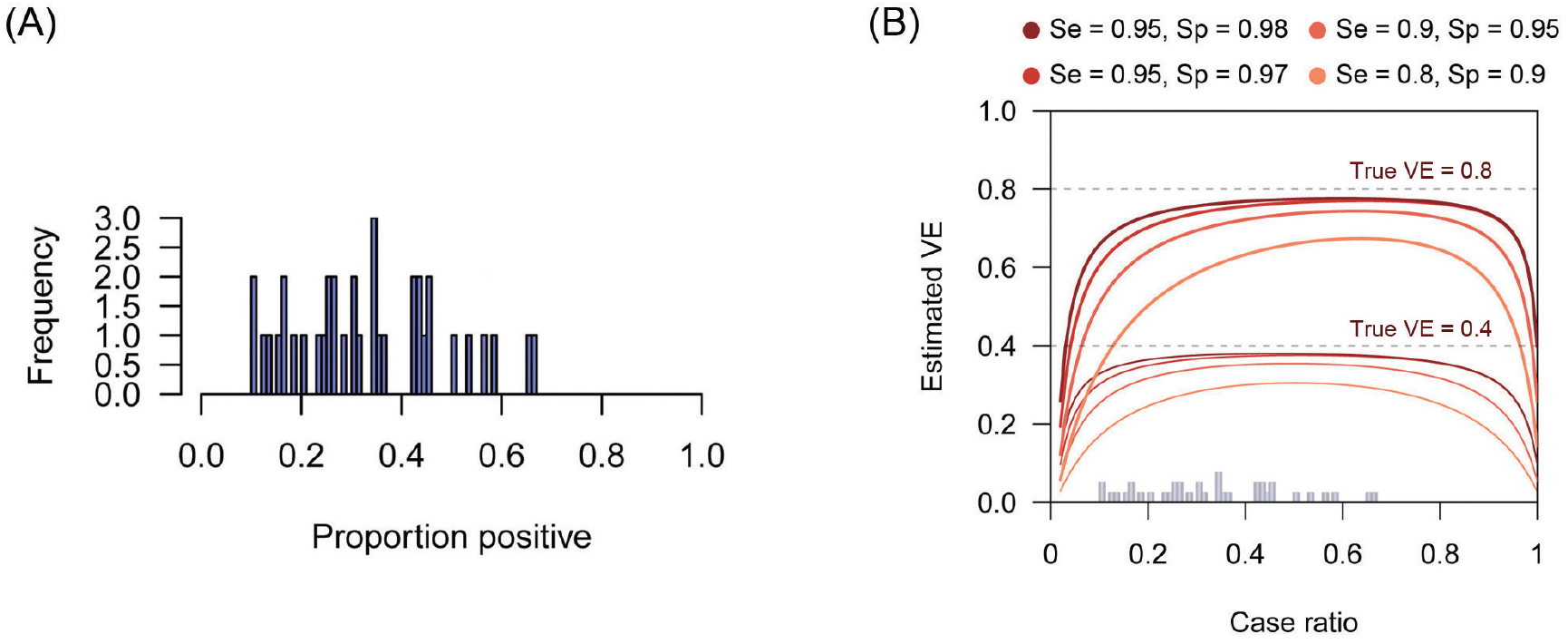
Biased VE estimates with varying case ratio and the observed proportion of positive patients. (A) The proportion of test positive patients in TND studies from systematic reviews. The proportions were retrieved from three systematic reviews [18, 19, 20]. (B) Estimated VE plotted against case ratio. Two sets of lines respectively correspond to different true VEs (80% and 40%, denoted by the dotted lines). The histogram in Panel (A) is overlaid on the x axis.

### 2.2 Bias correction in univariate analysis

#### 2.2.1 Model and statistical analysis

To develop a correction method that can address the bias presented in the previous section, we first model the case reporting process in the univariate setting as follows. Let us assume that incidence of TD and ND both follow Poisson-distributions. As presented in Section 2.1, the mean total incidence in the unvaccinated population is given as *λ*_*U*_ = (1 − *v*)*m*_*U*_ (*r*_1_*µ* + *r*_0_)*N*. Let 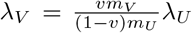 so that *λ*_*V*_ corresponds to the mean total incidence in the vaccinated population (= *Nv*) when *γ* = 1, i.e. VE=0. This definition is to ensure that parameters *γ* and *λ*_*V*_ are mutually independent. Let 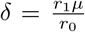 be the odds of the (medically-attended) target disease in the unvaccinated population. Using these four parameters *γ, δ, λ*_*V*_, *λ*_*U*_, we get the following table for (potentially mis-classified) mean case counts:

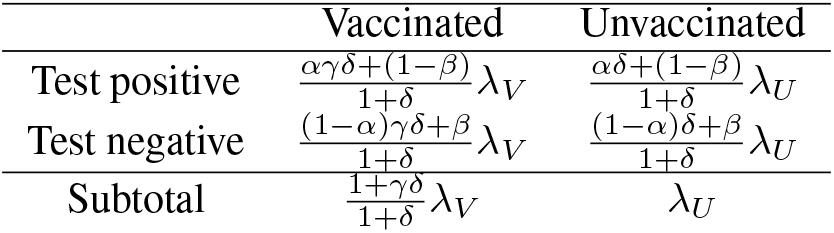

|  | Vaccinated | Unvaccinated |
| --- | --- | --- |
| Test positive | $\frac{\alpha\gamma\delta+(1-\beta)}{1+\delta}\lambda_V$ | $\frac{\alpha\delta+(1-\beta)}{1+\delta}\lambda_U$ |
| Test negative | $\frac{(1-\alpha)\gamma\delta+\beta}{1+\delta}\lambda_V$ | $\frac{(1-\alpha)\delta+\beta}{1+\delta}\lambda_U$ |
| Subtotal | $\frac{1+\gamma\delta}{1+\delta}\lambda_V$ | $\lambda_U$ |

When data *D* = (*X*_*V*_, *Y*_*V*_, *X*_*U*_, *Y*_*U*_) is obtained following this misclassified pattern, we can construct the likelihood of obtaining such data, given underlying parameters, as

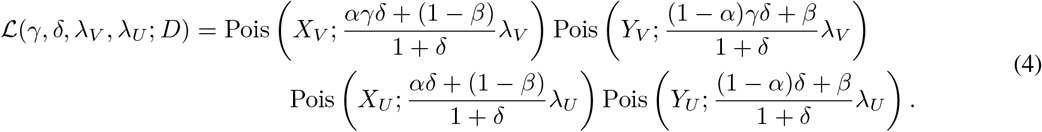

By maximising this likelihood over all parameters, we can obtain a maximum likelihood estimate (MLE) of the odds ratio *γ*^∗^ that accounts for misclassfication. Let us refer to

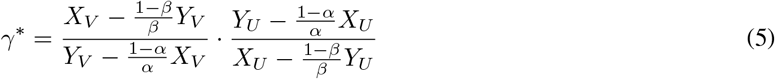

as the “corrected odds ratio”, which gives an unbiased estimate of *γ*. Comparing *γ*^∗^ with the the “raw” odds ratio 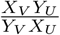, we find that the estimate can be corrected using the following substitution

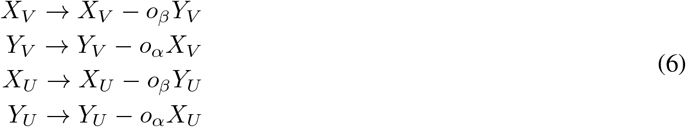

where 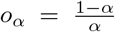 and 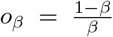 are the odds of diagnostic errors corresponding to sensitivity and specificity, respectively, which take 0 when sensitivity/specificity is perfect. Also note that the same odds ratio is obtained by taking the odds ratio of the reconstructed data table where the inverted classification matrix is applied:

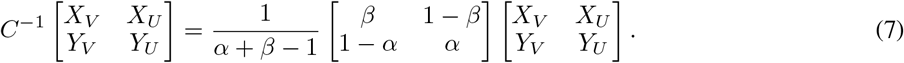

The determinant *c* = *α* + *β* − 1 is the Youden index of the test and satisfies 0 < *c* ≤ 1 (if *c* < 0, the test is not predictive and the definitions of positive/negative should be swapped).

In some (relatively rare) cases, one or more quantities in Eq. (6) may become negative due to random fluctuations in observation. Theoretically, negative values are not permitted in as true case counts, and thus such negative quantities would need to be truncated to 0. As a result, the corrected odds ratio can be either 0 or infinity. Such an estimate would suggest that the study does not have a sufficient sample size to properly evaluate VE and that the study design itself might need to be reconsidered.

The confidence interval for VE can be obtained by assuming log-normality of the odds ratio *γ*, i.e.,

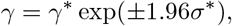

where *σ* is the shape parameter of the log-normal distribution and is empirically given as

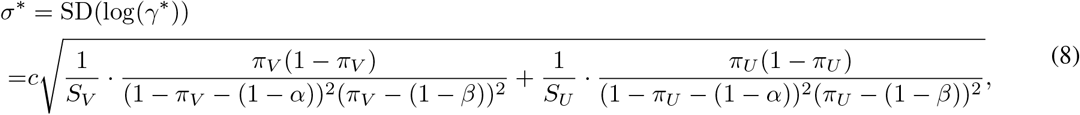

where *π*_*V*_ and *π*_*U*_ are observed (uncorrected) TD frequency (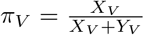 and 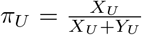). (See Appendix for details of the MLE and confidence intervals.)

#### 2.2.2 Simulation

To assess the performance of the corrected odds ratio given in Equation (22), we used simulation studies. TND study datasets were generated based on the likelihood presented in Equation (4), where the mean total sample size 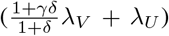 was set to be 3,000. Parameter values were chosen according to a range of scenarios shown in Table 1, and the true vaccine effectiveness VE = 1 − *γ* was compared with the estimates obtained from the simulated data. For each scenario, simulation was repeated 500 times to yield the distribution of estimates. Reproducible codes (including those for simulations in later sections) are reposited on GitHub (https://github.com/akira-endo/TND-biascorrection/).

**Table 1:**
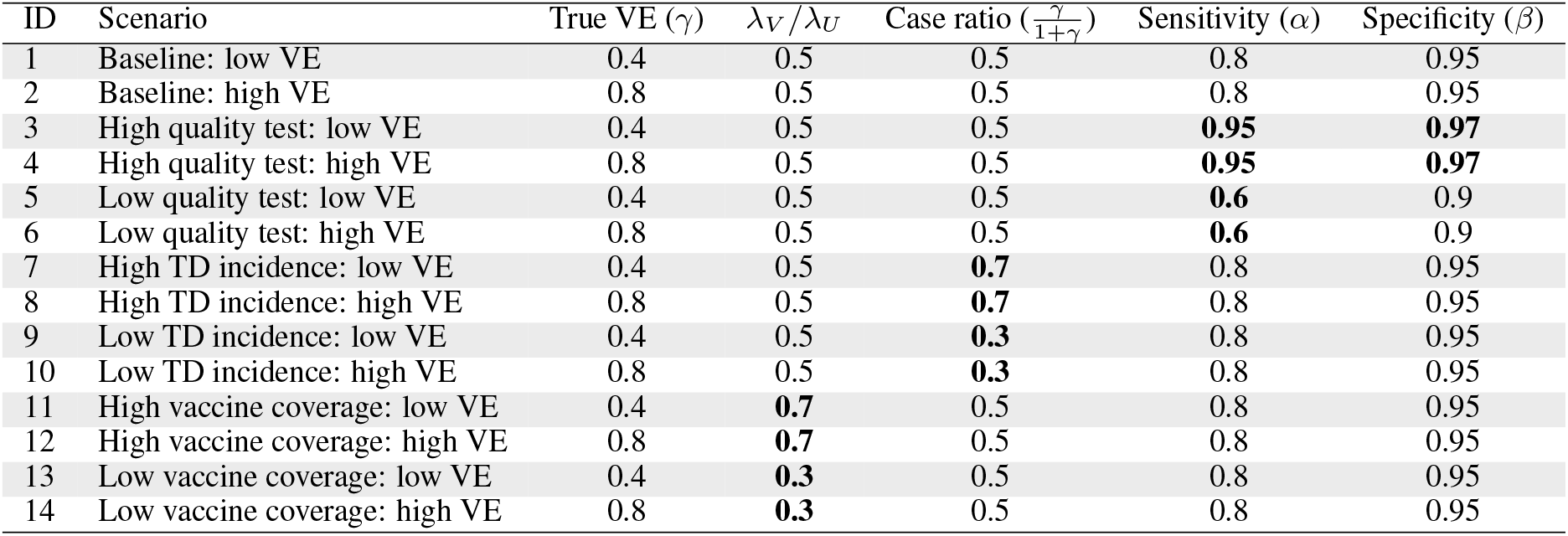
Simulation settings

| ID | Scenario | True VE ($\gamma$) | $\lambda_V/\lambda_U$ | Case ratio ($\frac{\gamma}{1+\gamma}$) | Sensitivity ($\alpha$) | Specificity ($\beta$) |
| --- | --- | --- | --- | --- | --- | --- |
| 1 | Baseline: low VE | 0.4 | 0.5 | 0.5 | 0.8 | 0.95 |
| 2 | Baseline: high VE | 0.8 | 0.5 | 0.5 | 0.8 | 0.95 |
| 3 | High quality test: low VE | 0.4 | 0.5 | 0.5 | **0.95** | **0.97** |
| 4 | High quality test: high VE | 0.8 | 0.5 | 0.5 | **0.95** | **0.97** |
| 5 | Low quality test: low VE | 0.4 | 0.5 | 0.5 | **0.6** | 0.9 |
| 6 | Low quality test: high VE | 0.8 | 0.5 | 0.5 | **0.6** | 0.9 |
| 7 | High TD incidence: low VE | 0.4 | 0.5 | **0.7** | 0.8 | 0.95 |
| 8 | High TD incidence: high VE | 0.8 | 0.5 | **0.7** | 0.8 | 0.95 |
| 9 | Low TD incidence: low VE | 0.4 | 0.5 | **0.3** | 0.8 | 0.95 |
| 10 | Low TD incidence: high VE | 0.8 | 0.5 | **0.3** | 0.8 | 0.95 |
| 11 | High vaccine coverage: low VE | 0.4 | **0.7** | 0.5 | 0.8 | 0.95 |
| 12 | High vaccine coverage: high VE | 0.8 | **0.7** | 0.5 | 0.8 | 0.95 |
| 13 | Low vaccine coverage: low VE | 0.4 | **0.3** | 0.5 | 0.8 | 0.95 |
| 14 | Low vaccine coverage: high VE | 0.8 | **0.3** | 0.5 | 0.8 | 0.95 |

We found that the uncorrected estimates, directly obtained from the raw case counts that were potentially misclassified, exhibited substantial underestimation of VE for most parameter values (Figure 3). On the other hand, our bias correction method was able to yield unbiased estimates in every setting, whose median almost correspond to the true VE. Although the corrected and uncorrected distributions were similar (with a difference in median ∼ 5%) when VE is relatively low (40%) and the test has sufficiently high sensitivity and specificity (95% and 97%, respectively), they became distinguishable with a higher VE (80%). With lower test performances, the bias in the VE estimates can be up to 10-20%, which may be beyond the level of acceptance in VE studies.

**Figure 3:**
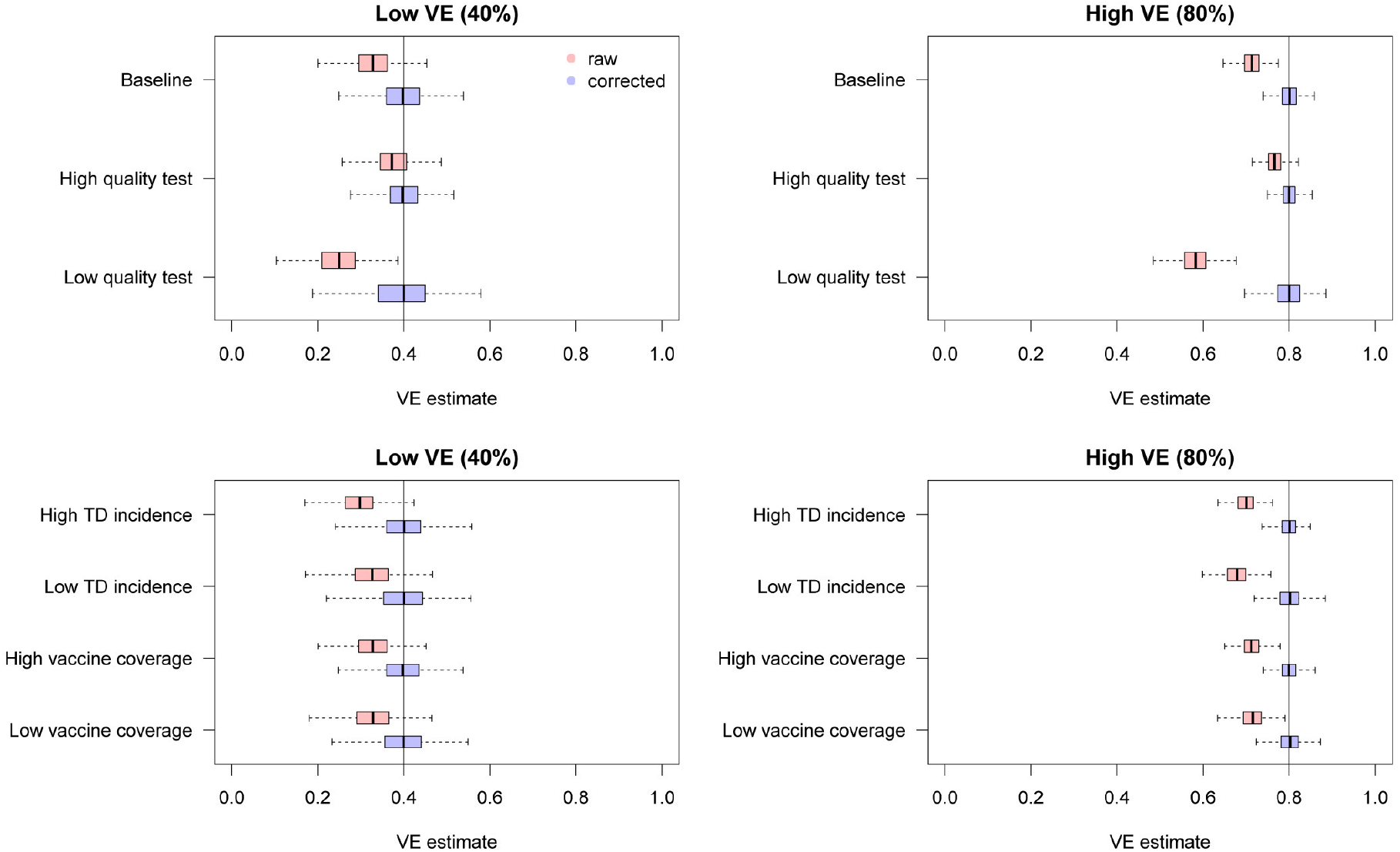
Bias correction for simulated data in the univariate setting. The distributions of bias-corrected VE estimates (boxplots in blue) are compared with those of raw VE estimates without correction (red). Five hundred independent datasets were randomly generated for each set of parameter values, and the corrected and uncorrected VE estimates are compared with the true value (black solid line).

#### 2.2.3 Bias correction of VEs reported in previous studies

We have seen that the degree of bias for uncorrected VE estimates depends on parameter values. To explore the possible degree of bias in existing VE studies, we extracted the reported crude VEs (i.e. VEs without adjustments of potential confounders) from two systematic reviews [18, 20] (Young et al. [19] was not included because they did not report case counts) and applied our bias correction method assuming different levels of test sensitivity and specificity. The case counts for each study summarised in the reviews were considered eligible for the analysis if the total sample size exceeded 200. Varying the assumed sensitivity and specificity, we investigated the possible discrepancy between the reported VE (or crude VE derived from the case counts if unreported in the reviews) and bias-corrected VE. We did not consider correcting adjusted VEs because it requires access to the original datasets.

Figure 4 displays the discrepancy between the reported VE and bias corrected VE corresponding to a range of assumptions on the test performance. Many of the extracted studies employed polymerase chain reaction (PCR) for the diagnostic test, which is expected to have a high performance. However, the true performance of PCR cannot be definitively measured as there is currently no other gold-standard test available. Figure 4B suggests that even a slight decline in the test performance can introduce a non-negligible bias in some parameter settings. Our bias correction methods may therefore also be useful in TND studies using PCR, which would enable a sensitivity analysis accounting for potential misdiagnosis by PCR tests. In this light, it is useful that the corrected odds ratio 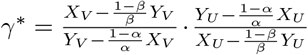 is a monotonic function of both *α* and *β* (given that all the four components are positive). The possible range of VE in a sensitivity analysis is obtained by supplying *γ*^∗^ with the assumed upper and lower limits of sensitivity and specificity.

**Figure 4:**
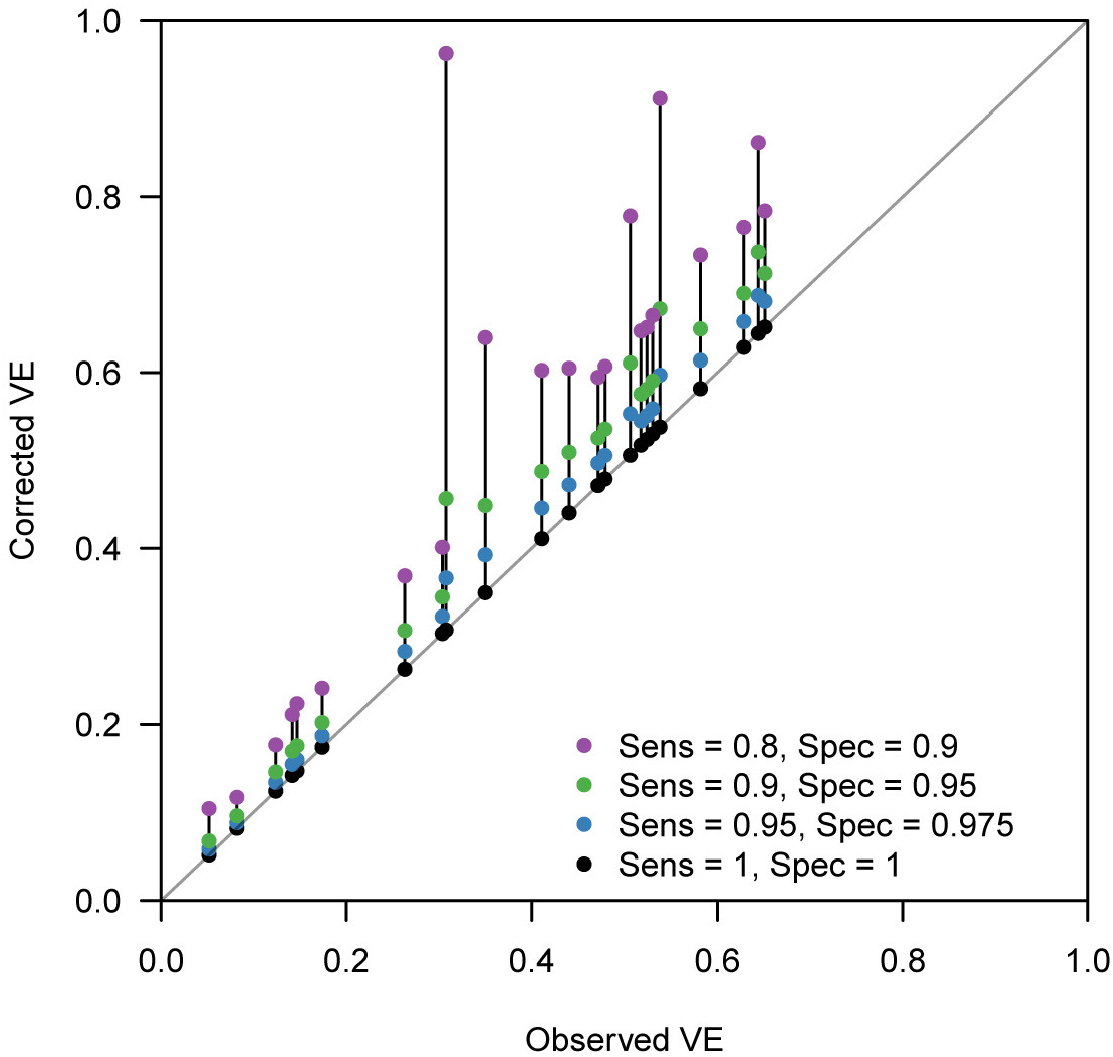
Bias correction method applied to published VE estimates assuming various test sensitivity and specificity. Case count data were extracted from two systematic reviews [18, 20]. Each connected set of dots show how (crude) VE estimates reported in the review varies when imperfect sensitivity and specificity are assumed. Black dots on the grey diagnoal line denote the original VEs reported in the reviews (where sensitivity = specificity = 1) and coloured dots show the estimated VE considering potential misclassification.

### 2.3 Bias correction in multivariate analysis

#### 2.3.1 Theoretical framework

TND studies often employ a multivariate regression framework to address potential confounding variables such as age. The most widespread approach is to use linear models (e.g., logistic regression) and include vaccination history as well as other confounding variables as covariates. The estimated linear coefficient for vaccination history can then be converted VE (in the logistic regression model, the linear coefficient for vaccination history corresponds to log(1 − VE)).

In this situation, the likelihood function now reflects a regression model and thus the bias-corrected estimate in the univariate analysis (Equation (5)) is no longer applicable. We therefore need to develop a separate multivariate TND study framework to correct for bias in multivariate analysis. Suppose that covariates *ξ* = (*ξ*^1^, *ξ*^2^, …, *ξ*^*n*^) are included in the model, and that *ξ*^1^ corresponds to vaccination history (1: vaccinated, 0: unvaccinated). *ξ* is expected to have a certain distribution over the total population *N*, and let us denote the frequency density of covariates *ξ* by ∫ *N* (*ξ*), where *N* (*ξ*)*dξ* = *N*. Let *ρ*_1_(*ξ*) and *ρ*_0_(*ξ*) be the conditional probabilities that an individual is included in the study with TD and ND, respectively, given covariates *ξ*. Incorporating misclassification, the probability of an individual *i* with covariates *ξ*_*i*_ being included and tested positive/negative will be

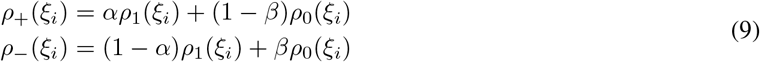

Assuming that disease incidences follow Poisson distributions, as in the univariate case, we can obtain the probability density of observing data *D* = {*Z*_*i*_, *ξ*_*i*_}_*i*=1,2,…*S*_ (*Z*_*i*_ denotes the test result of individual *i*) as

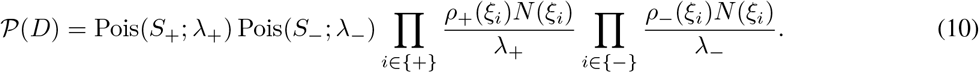

where *λ*_+_ and *λ*_−_ are the mean incidence of being included in the study and tested positive/negative: *λ*_±_ = ∫ *ρ*_±_ (*ξ*)*N* (*ξ*)*dξ*. The first two Poisson distributions on the right-hand side of Eq. (10) give the probability that the study yields *S*_+_ positive and *S*_-_ negative subjects. The products that follow represent the probability density for covariates *ξ*_*i*_ observed in the positive/negative group.

Suppose that we model this system using a parameter set *θ*. We could directly model *ρ*_1_(*ξ*_*i*_; *θ*) and *ρ*_0_(*ξ*_*i*_; *θ*); however, it is often more convenient to model the binomial probability for the true outcome 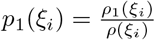 and 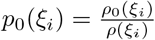, where *ρ*(*ξ*_*i*_) = *ρ*_1_(*ξ*_*i*_) + *ρ*_0_(*ξ*_*i*_) = *ρ*_+_(*ξ*_*i*_) + *ρ* (*ξ*_*i*_) is the probability density of being included in the study given covariates *ξ*, because the absolute scale of incidence is rarely of a primary concern. The binomial probabilities for the respective observed outcomes (with errors) are then given by:

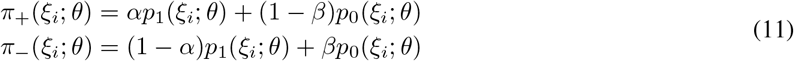

Let us use parameter set *θ* to model the binomial probabilities *π*_+_ (and *π*_−_) and assume that another set of parameters *η* (nuisance parameters) characterise *ρ*(*ξ*_*i*_). Then our objective is reduced to the estimation of *θ* and *η*.

Rearranging Equation (10), we get the joint likelihood for *θ* and *η*:

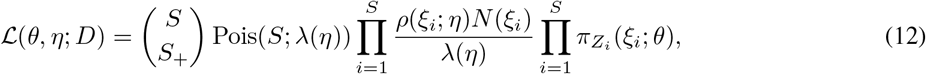

where *λ*(*η*) is the overall mean incidence: *λ*(*η*) = ∫ *ρ*(*ξ*; *η*)*N* (*ξ*)*dξ*. The factor outside the products on the right-hand side of Eq. (12) is the probability that the study yields *S* subjects of which *S*_+_ are positives and *S*_−_ are negatives. The first product is the probability density for covariates *ξ*_*i*_ observed in data *D*, and the second product is the binomial probabilities for the test results *Z*_*i*_. When only *θ* is of our concern, we can obtain the MLE for *θ* by maximising

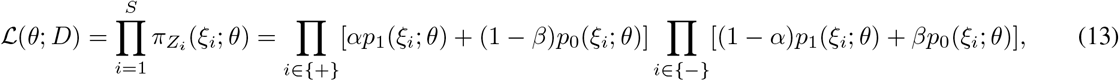

as *θ* and *η* are separate in the likelihood (12). With the estimate *θ*^∗^, the VE estimate for an individual with covariates *ξ*^2:*n*^ = (*ξ*^2^, *ξ*^3^, …, *ξ*^*n*^) is given as (1 - odds ratio):

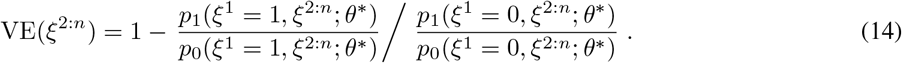

#### 2.3.2 Direct likelihood method for the logistic regression model

The logistic regression model is well-suited for modelling binomial probabilities *p*_1_ and *p*_0_. The log-odds 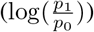 is characterised by a linear predictor as:

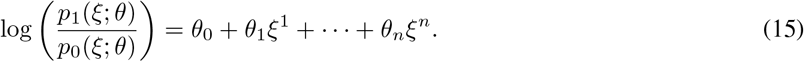

In the logistic regression model where covariate *ξ*_1_ indicates vaccination history, the corresponding coefficient *θ*_1_ gives the VE estimate: VE = 1 − exp(*θ*_1_). Due to the assumed linearity, the estimated VE value is common across individuals regardless of covariates *ξ*^2:*n*^.

We can employ the direct likelihood method by combining Equations (13) and (15). The usual logistic regression optimises *θ* by assuming that the test results follow Bernouli distributions *Z*_*i*_ ∼ Bernouli(*p*_1_(*ξ*_*i*_; *θ*)) (*Z*_*i*_ = 1 for positive test results and 0 for negative). To correct the misclassification bias, we instead need to use the modified probabilities given by Eq. (11) to construct the likelihood accounting for diagnostic error, i.e.,

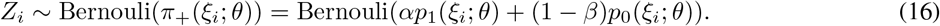

Parameter *θ* is estimated by directly maximising the probability of observing {*Z*_*i*_} based on Eq. (16)

Note that as long as the binomial probability is the modelling target, other type of models (e.g. probit model) could also be employed under a similar framework.

#### 2.3.3 Multiple overimputation method to be combined with existing tools

The direct likelihood method presented in the previous section is the most rigorous MLE approach and would therefore be preferable whenever possible. However, it is often technically-demanding to implement such approaches as it involves re-defining the likelihood; if we wanted to use existing tools for logistic regression (or other models), for example, we would need to modify the internal algorithm specifying the likelihood computation. To ensure that researchers are able to employ correction methods without reprogramming the underlying software algorithms, we also propose another method, which employs a multiple overimputation (MO) framework [12] to account for misclassification. Whereas multiple imputation only considers missing values, MO is proposed as a more general concept which includes overwriting mismeasured values in the dataset by imputation. In our multivariate bias correction method, test results in the dataset (which are potentially misclassified) are randomly overimputed.

Let *M* be an existing estimation software tool whose likelihood specification cannot be reprogrammed. Given data *d* = {*z*_*i*_, *ξ*_*i*_}_*i*=1,2,…*S*_, where *z*_*i*_ denotes the true disease state (*z* = 1 for TD and *z* = 0 for ND), *M* would be expected to return at least the following two elements: the point estimate of VE (*ε*_*d*_) and the predicted binomial probability 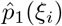 for each individual *i*. From the original observed dataset *D*, imputed datasets 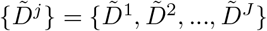 are generated by the following procedure.

1. For *i* = 1, 2, …, *S*, impute disease state 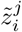 based on the test result *Z*_*i*_. Each 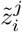 is sampled from a Bernouli distribution conditional to *Z*_*i*_:

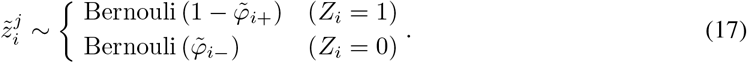 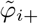 and 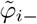 are estimated probabilities that the test result for individual *i* is incorrect (i.e., *z*_*i*_ ≠ = *Z*_*i*_) given *Z*_*i*_. The sampling procedure (17) is therefore interpreted as the test result *Z*_*i*_ being “flipped” at a probability 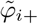 or 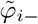. Later we will discuss possible procedures to obtain these probabilities.
2. Apply *M* to 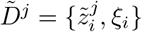 to yield a point estimate of VE (*ε*^*j*^).
3. Repeat 1., 2. for *j* = 1, 2, …, *J* to yield MO estimates {*ε*^*j*^}_*j*=1,…*J*_.

Once MO estimates {*ε*^*j*^} are obtained, the pooled estimate and confidence intervals of VE are obtained by appropriate summary statistics, e.g., Rubin’s rules. As long as the estimated “flipping” probabilities 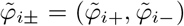 are well chosen, this MO procedure should provide an unbiased estimate of VE with a sufficiently large number of iterations *J*.

There can be multiple candidates for the flipping probability estimate 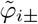. Here we discuss two possible options: parametric bootstrapping and the expectation-maximisation (EM) algorithm. Our simulation showed that parametric bootstrapping is preferable (see the supplementary document).

##### (i) Parametric bootstrapping

The simplest option to estimate 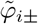 is to use Bayesian probability

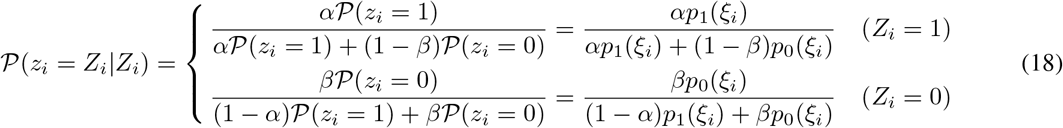

The true binomial probabilities *p*_0_(*ξ*_*i*_), *p*_1_(*ξ*_*i*_) are not known, but their estimators are derived with the inverted classification matrix in the same manner as Eq. (7). By substituting 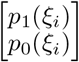 with 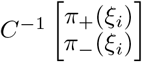, we get

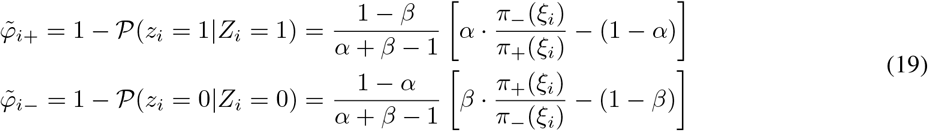

These probabilities can be computed provided the odds of the test results 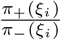. We employ a parametric approach and approximate this odds by applying estimation tool *M* to the original data *D*; i.e., the predicted binomial probability 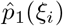 obtained from *D* is used as a proxy of *π*_+_(*ξ*_*i*_). Generally it is not assured that true and observed probabilities *p*_1_(*ξ*_*i*_) and *π*_+_(*ξ*_*i*_) have the same mechanistic structure captured by *M* ; however, when our concern is limited to the use of model-predicted probabilities to smooth the data *D*, we may expect for *M* to provide a sufficiently good approximation with realistic test sensitivity and specificity. The above framework can be regarded as a variant of parametric bootstrapping methods as MO datasets are generated from data *D* assuming a parametric model *M*. The whole bias correction procedure is presented in pseudocode (Algorithm 1); a sample R code is also available on GitHub https://github.com/akira-endo/TND-biascorrection/).

###### Algorithm 1 Multiple imputation with parametric bootstrapping

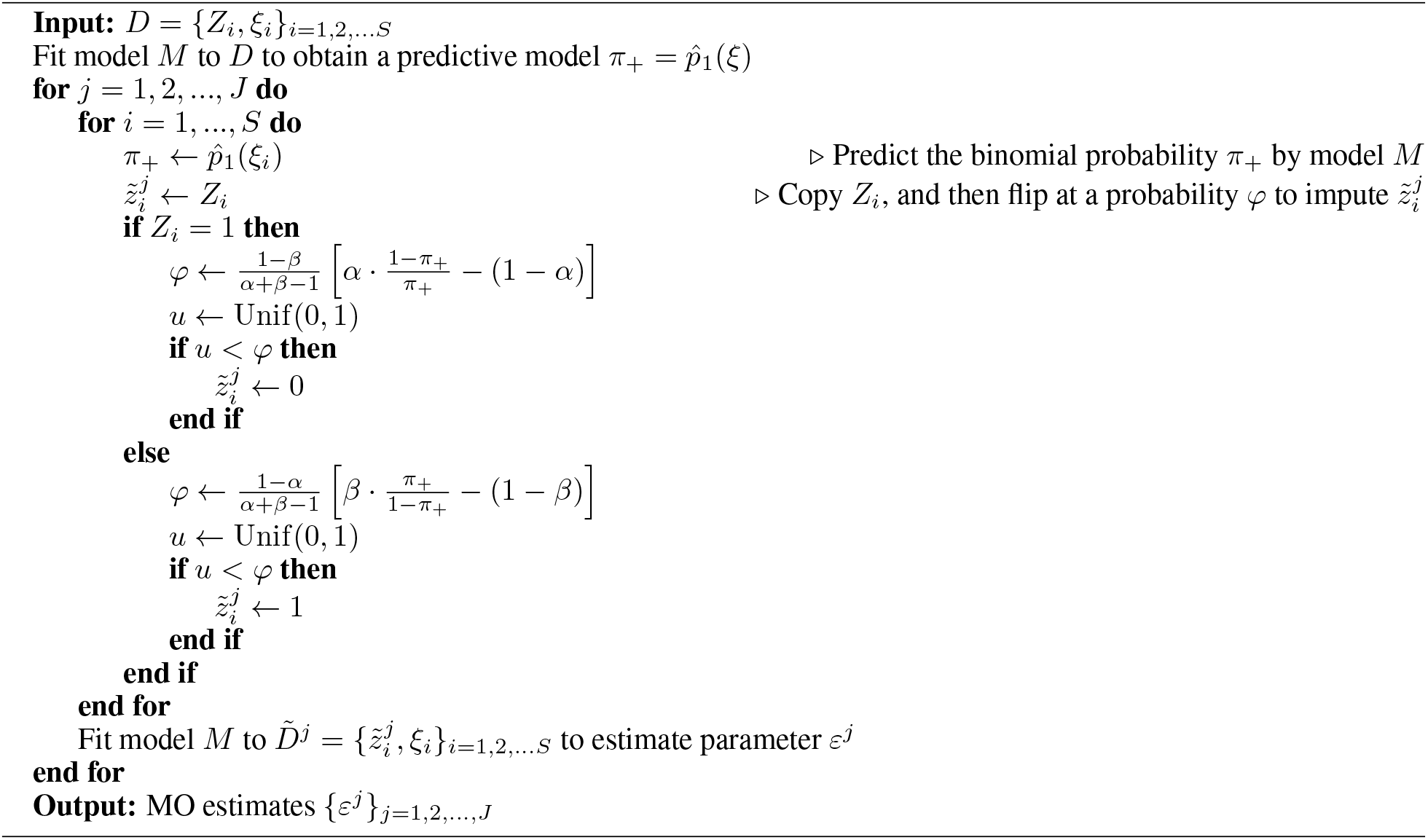

##### (ii) EM algorithm

Another possible approach is to use EM algorithm as proposed by Magder et al. [21], where 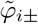 can be estimated by iterations (see the supplementary document for details). However, in our simulation we found that the performance of EM algorithm was inferior to the other two alternatives (direct likelihood and parametric bootstrapping). The three methods all provided effectively identical distributions of estimates in most settings, but in some settings the EM algorithm produced extreme estimates (VE < 0 or > 1) slightly more often than the other two. We therefore recommend parametric bootstrapping as the first choice of bias correction method when the direct likelihood approach is inconvenient.

#### 2.3.4 Simulation of bias correction with parametric bootstrapping

To assess the performance of this method, we used the same simulation framework as in the univariate analysis (Table 1). In addition to vaccination history (denoted by *ξ*^1^), we consider one categorical and one continuous covariate. Let us assume that *ξ*_2_ represents the age group (categorical; 1: child, 0: adult) and *ξ*_3_ the pre-infection antibody titre against TD (continuous). Suppose that the population ratio between children and adults is 1:2, and that *ξ*_3_ is scaled so that it is standard normally distributed in the population. For simplicity, we assumed that all the covariates are mutually independent with regard to the distribution and effects (i.e., no association between covariates and no interaction effects). The relative risk of children was set to be 2 and 1.5 for TD and ND, respectively, and a unit increase in the antibody titre was assumed to halve the risk of TD (and not to affect the risk of ND). The mean total sample size *λ* was set to be 3,000, and 500 sets of simulation data were generated for each scenario. VE estimates were corrected by the parametric bootstrapping approach (the number of iterations *J* = 100) and were compared with the raw (uncorrected) VE estimates.

Figure 5 shows the distributions of estimates with and without bias correction in the multivariate setting. Our bias correction (parametric bootstrapping) provided unbiased estimates for all the scenarios considered. Overall, biases in the uncorrected estimates were larger than those in the univariate setting. In some scenarios, the standard error of the bias-corrected estimates was extremely wide. This was not because of the failure of bias correction, but because of the uncertainty already introduced before misclassification. The standard errors in those scenarios were very large even with perfect test sensitivity and specificity as can be seen in Figure 6. Larger sample size is required to yield accurate estimates in those settings, as the information loss due to misclassification will be added on top of the inherent uncertainty in the true data.

**Figure 5:**
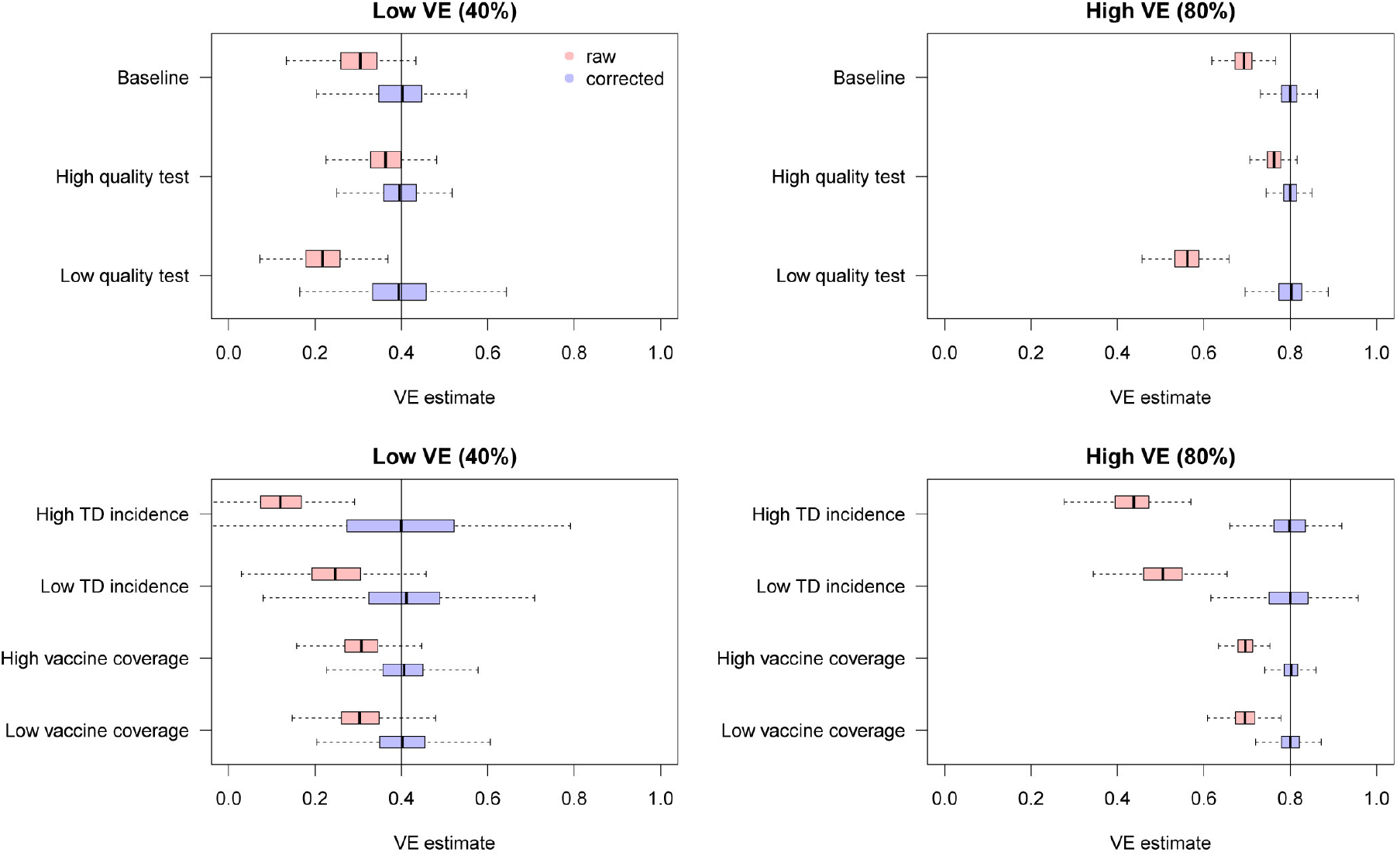
Bias correction for simulated data in the multivariate setting. The distributions of bias-corrected (blue) and uncorrected (red) VE estimates from 500 simulations are compared. Dotted lines denote median and black solid lines denote the true VE. The parametric bootstrapping bias correction method was used for bias correction.

**Figure 6:**
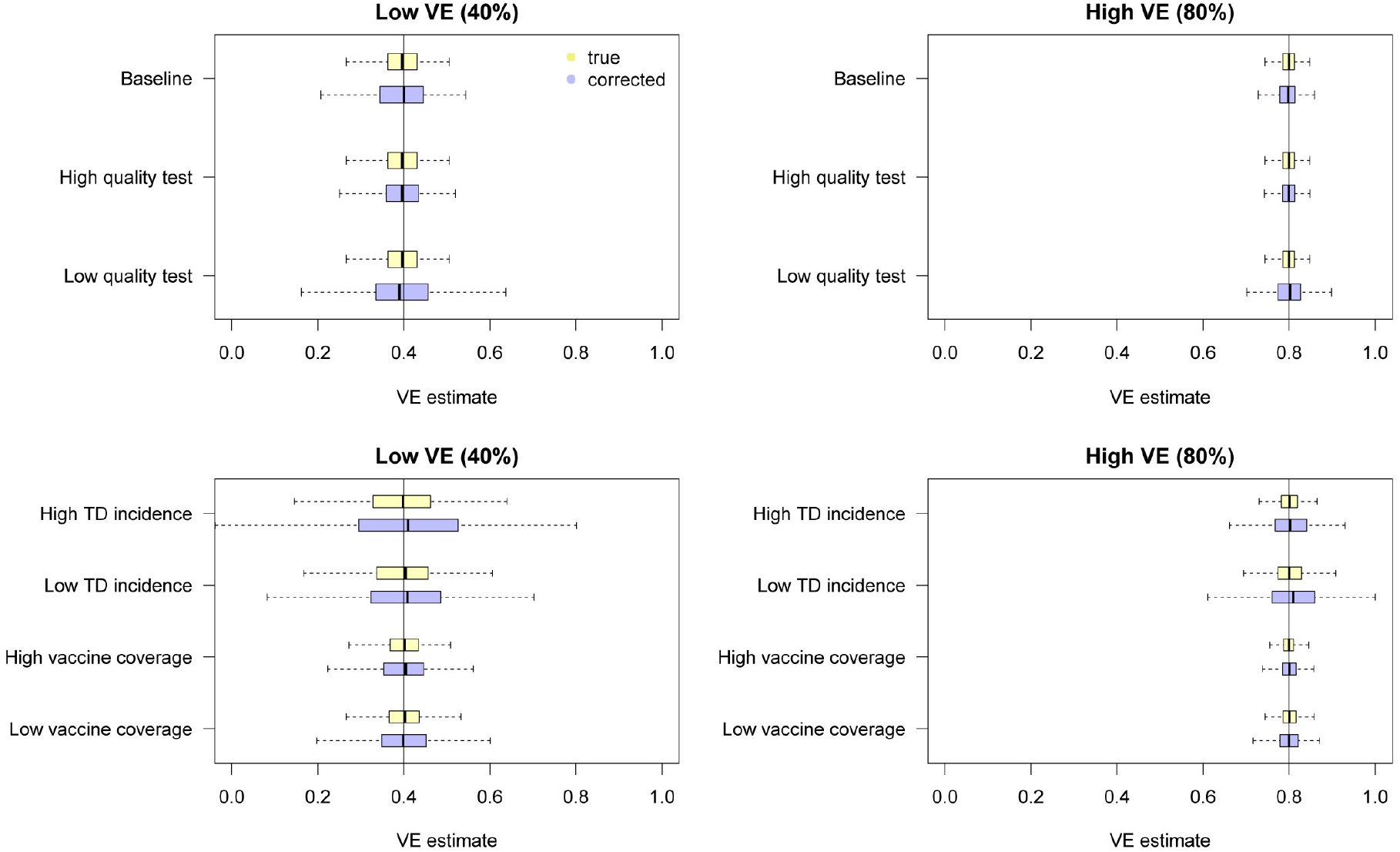
Uncertainty in VE estimates obtained from the true/misclassified datasets in the multivariate setting. The distributions of VE estimates from the simulated true (yellow) and misclassified (light blue) data are shown. The direct likelihood method was employed to correct biases in the misclassified data.

#### 2.3.5 Increased uncertainty introduced by misclassification

Although our bias correction methods provide unbiased VE estimates from potentially misclassified test results, the resulting uncertainty is larger than that which would be obtained from estimates using the true disease status. In Figure 6, we compared bias-corrected estimates obtained from misclassified data (by the direct likelihood method) with those obtained from the true data (i.e., 100% sensitivity and specificity). Although both estimates are unbiased around the true value, the results from the misclassified data exhibit higher variability (by a factor of 1.1-3.0) due to the loss of information caused by misdiagnosis. Increased uncertainty due to misclassification should be carefully considered when one calculates the power of test-negative design studies. Overestimated test performance may not only underestimate the true VE but also lead to overconfidence.

### 2.3.6 The number of confounding variables

We investigated how the bias in uncorrected VE estimates can be affected by the number of confounding variables. In addition to the vaccine history *ξ*^1^, we added a set of categorical/continuous confounding variables to the model and assessed the degree of bias caused by misclassification. The characteristics of the variables were inherited from those in Section 2.3.4: categorical variable “age” and continuous variable “pre-infection antibody titre”. That is, individuals were assigned multiple covariates (e.g., “categorical variable A”, “categorical variable B”, …, “continuous variable A”, “continuous variable B”, …) whose distribution and effect were identical to “age” (for categorical variables) and “antibody titre” (for continuous variables) in Section 2.3.4. No interaction between covariates was assumed. The covariate set in Section 2.3.4 being baseline (the number of covariates: (vaccine, categorical, continuous) = (1, 1, 1)), we employed two more scenarios with a larger number of covariates: (1, 3, 3) and (1, 5, 5).

The simulation results are presented in Figure 7. Overall, additional confounding variables led to more severe bias in the uncorrected VE estimates towards underestimation. As shown in Figure 2A, the degree of bias is strongly affected by the case ratio: the ratio between the risk of TD and ND. More confounding variables in a population result in higher heterogeneity in individuals’ risk of TD and ND. This may account for the association between the degree of bias and the number of confounding variables; more individuals in a highly heterogeneous population may fall in the outer range of the case ratio shown in Figure 2A, substantially contributing to the misclassification bias.

**Figure 7:**
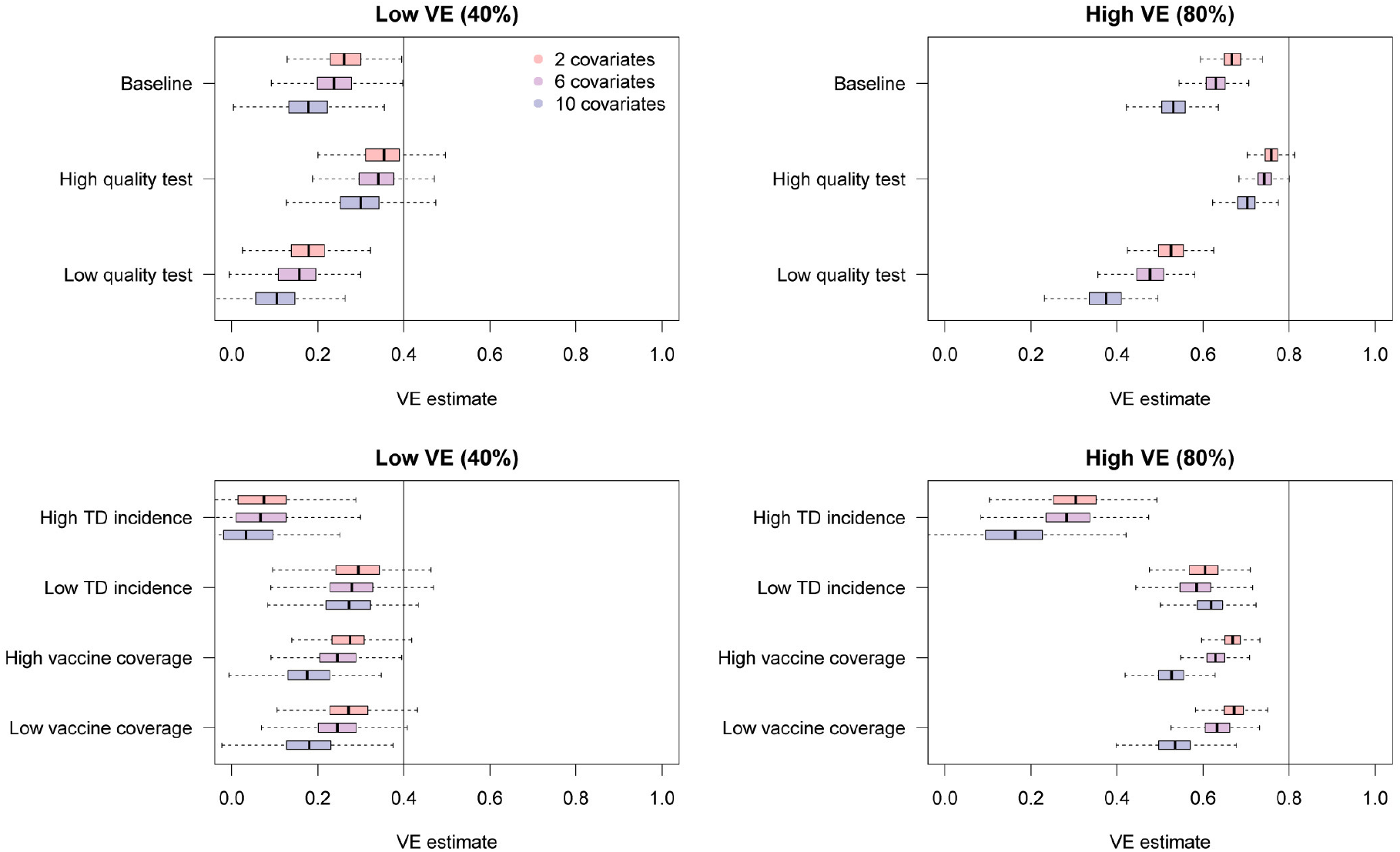
Bias in raw VE estimates from simulated data in the presence of different numbers of confounding variables. The distributions in red, purple and blue correspond to uncorrected VE estimates in the presence of 2, 6 and 10 confounding variables in addition to the vaccination history.

## 3 Discussion

Misclassification caused by imperfect diagnostic tests can potentially lead to substantial biases in TND studies. By considering the processes involved in VE estimation, we have characterised the degree of bias potentially caused by diagnostic misclassification in different parameter settings, finding that VE can be noticeably underestimated, particularly when the ratio between TD and ND cases in the study data is unbalanced. To address this potential bias, we developed multiple bias correction methods that address test misclassification and provide unbiased VE estimates in both univariate and multivariate settings. When the test sensitivity and specificity are known or assumed, those values can be used to restore the true VE estimate by a relatively simple statistical procedure. Using simulations, we showed that our methods could successfully eliminate the bias in VE estimates obtained from misclassified data, although some uncertainty was introduced as a result of the information loss.

We believe that our methods could therefore enable researchers to address possible misclassification in their data and report unbiased VE estimates even when imperfect tests had to be used. Such methods could also help in the scaling up of TND studies, as tests with limited performance are usually inexpensive and logistically convenient. Even when high-performance diagnostic tests including PCR techniques are available, the risk of misdiagnosis may not be negligible in certain settings, and our methods could further be used to perform sensitivity analysis to properly address the possibility of bias in such cases.

Although TND is a relatively new study design, first appearing in a publication in 2005 [22], it has gained broad popularity and is becoming a standard approach in VE studies. TND is believed to minimise the bias caused by different health seeking behaviour of individuals, but one of the largest factors that have contributed to its widespread use is the fact that data collection can be completed within clinical setups [1]. Whereas cohort or case-control studies usually require additional efforts including follow up or recruitment of non-patients, TND studies only involve patients visiting healthcare facilities with suspects of certain diseases and thus routinely collected clinical data can be easily adapted for analysis. For diseases of which suspected patients routinely undergo lab tests, TND may be one of the most convenient options to generate epidemiological insights into the effect of specific prevention/treatment. VE studies of influenza, for which TND is most frequently used, often use PCR as a diagnostic tool for better data quality [20]. However, such studies usually involve intensive effort and cost, and thus may only be feasible by large-scale research bodies. Our bias correction methods may open a possibility of wider use of clinical data, which could potentially provide rich epidemiological insights, especially in settings where rapid tests are routinely used for diagnosis. For example, rapid influenza diagnostic tests are routinely used for inpatient clinics and hospitals in Japan, and such clinical data have facilitated a number of TND studies [23, 24, 25, 26, 27, 28]. Such studies based on rapid tests could benefit from our methods, as it would provide strong support for the validity of their estimates. Our methods may also be useful in resource limited settings or for diseases without high-performance diagnostic tools. Even in resourceful settings where high-performance tests are available, the slight possibility of misclassification might not always be neglected. Although PCR tests are currently used as a gold-standard for influenza diagnosis, their sensitivity and specificity may not be exact 100%; especially, the sensitivity of the test depends not only on microbiological technique but also on the quality of swab samples from patients. In addition, it is suggested that the sensitivity of PCR tests may change during the time course of infection [29] and be sufficiently high only during a limited time window. Our simulation study also indicated that a high heterogeneity in individual characteristics in study samples might increase the bias in the VE estimate. Our methods could enable researchers to implement sensitivity analysis by assuming the possible test sensitivity and specificity in such cases.

Our bias correction methods are also intended to be reasonably straightforward for researchers to introduce. Existing estimation tools including software libraries and packages are often used in epidemiological analyses, and most of them have specific requirements for data input and output. Modifying the procedure employed by such tools in a way which is unexpected by the authors is usually impossible or requires advanced technical skills including reprogramming of the underlying algorithms. Incorporating the MO approach, our parametric bootstrapping bias correction method only involves data manipulation and does not require modification of the estimation algorithm. The only inputs required to produce MO datasets are the assumed test sensitivity and specificity (*α, β*) and the model-predicted binomial probability for the test result (*π*_+_) for each individual. Once multiple sets of data are randomly generated, any type of analysis can be performed as long as they produce numerical estimates to be summarised over the MO datasets. Of particular note is that our methods for multivariate analysis (including the direct likelihood method) allow stratification of sensitivity and specificity among individuals. Therefore, the users can employ more complex misclassification mechanisms including time-varying test performance or test performance affected by individual characteristics. Datasets with a mixture of different diagnostic tools [3, 30] can also be handled by applying different values for each test.

There are some limitations to our study. We only focused on misclassification of diagnosis (i.e., misclassified outcomes) and did not consider potential misclassification of covariates (e.g., vaccine history and other confounding variables), which is another important type of misclassification in TND studies [10]. Further, it is generally not easy to plausibly estimate the sensitivity and specificity for measurement of covariates (e.g. recall bias), which must be known or assumed to implement bias correction. However, if reliable estimates are available, an extension of our approach may yield bias-corrected VE estimates in the presence of covariate misclassification. Such consideration remains to be discussed in future work. Moreover, to keep our focus only on diagnostic misclassification, our methods rested on the assumption that other sources of bias in TND studies are nonexistent or properly addressed. Potential sources of bias in TND studies have been discussed elsewhere [13, 31], and the researchers conducting TND studies need to carefully consider the possibility of such biases in addition to the diagnostic misclassification. Lastly, it must be noted that our methods depend on the assumed test sensitivity and specificity, and that misspecifying those values can result in an improper correction. The sensitivity and specificity of tests are usually reported by manufacturers in a comparison of the test results with gold-standard tests; however, when such gold-standard tests themselves are not fully reliable or when no available test has satisfactory performance to be regarded as gold-standard, specifying sensitivity and specificity of a test is in principle impossible. Further, test performances reported by manufacturers might lack sufficient sample size or might not be identical to those in the actual study settings. Use of composite reference standards [32, 33] or external/internal validation approaches [34] may help overcome these problems.

Although the presence of imperfect diagnosis limits the quality of clinical data, data with such uncertainty can still hold useful information, and this information can be transformed into useful insights by appropriate statistical processing. Our bias correction methods were developed primarily for TND studies, but a similar approach could be applied to broader classes of estimation problems with misclassification. The value of routinely collected data in healthcare settings has become widely recognised with the advancement of data infrastructure, and we believe our methods could help support the effective use of such data.

## 4 Conclusion

Bias correction methods for the test-negative design studies were developed to address potential misclassification bias due to imperfect tests.

## Data Availability

The study used simulated data only. The code used to generate the data is reposited online (https://github.com/akira-endo/TND-biascorrection/)

## Abbreviations

VE: vaccine effectiveness
TND: test-negative design
TD: target disease
ND: non-target disease
PCR: polymerase chain reaction
MLE: maximum likelihood estimate
MO: multiple overimputation
EM: expectation-maximisation.

## Appendix

### Maximum likelihood estimates and confidence intervals in the univariate setting

Expanding Equation (4) in the main text, we get

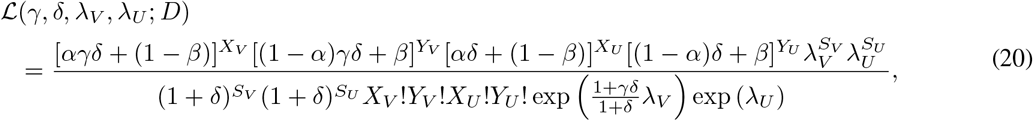

where *S*_*V*_ = *X*_*V*_ + *Y*_*V*_ and *S*_*U*_ = *X*_*U*_ + *Y*_*U*_.

For mathematical convenience, we change the variable *λ*_*V*_ to 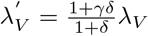. Let 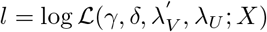. Partial derivatives of *l* are

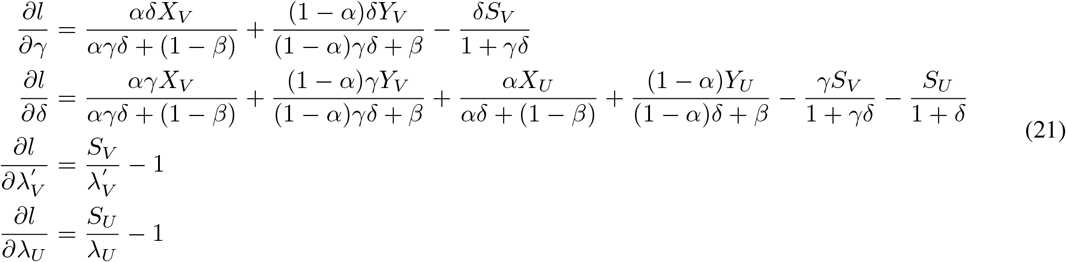

Equation (21) gives the maximum likelihood estimates:

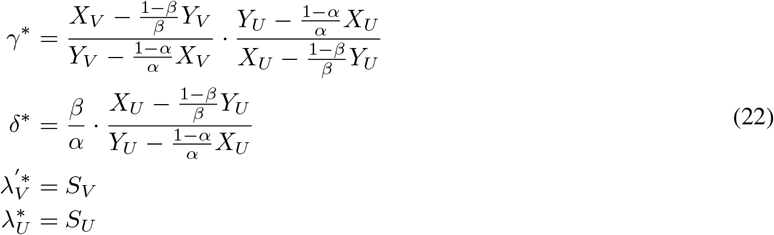

The confidence intervals for parameters can be constructed using the Fisher’s information matrix from Equation (21). 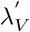 and *λ*_*U*_ are independent from other parameters and

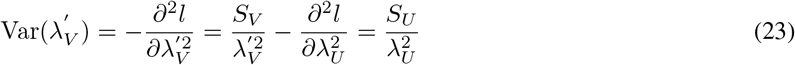

We log-transform *γ* and *δ* for mathematical convenience. Noting 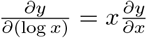, we get

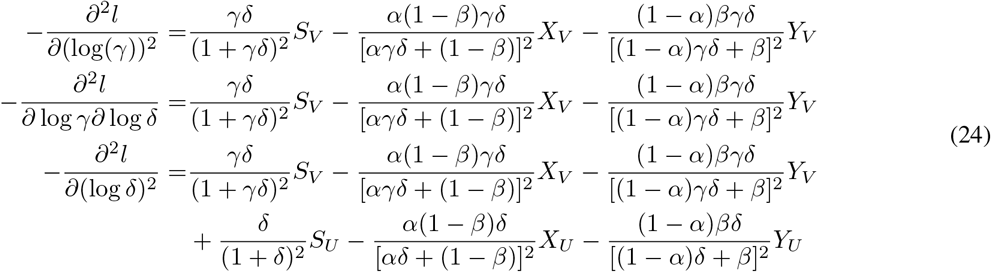

With the parameter values estimated in Eq. (22), we get the following information matrix

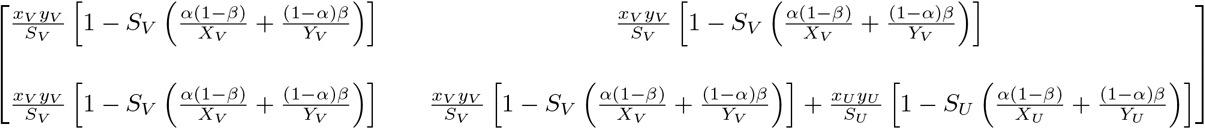

where 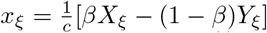 and 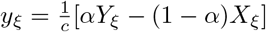 are the true case counts (without misclassification) for *ξ* = *V, U*. Let *p*_*V*_ = *x*_*V*_ */*(*x*_*V*_ + *y*_*V*_) and *p*_*U*_ = *x*_*U*_ */*(*x*_*U*_ + *y*_*U*_) be the corresponding true binomial probabilities.

The inverse of the information matrix provides variance of estimates: in particular, for log *γ* we get

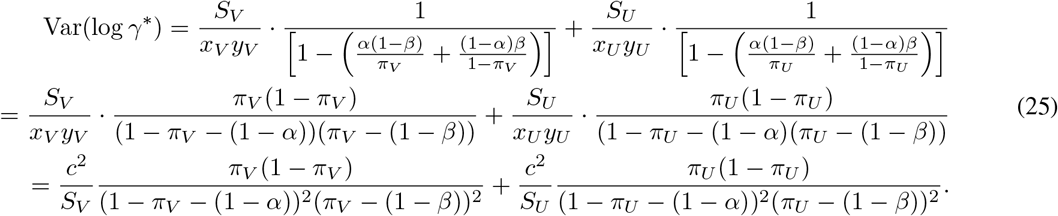

We can relate this to the true standard error that would be obtained with perfect tests,

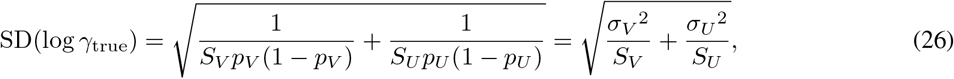

or to the observed standard error (without correction),

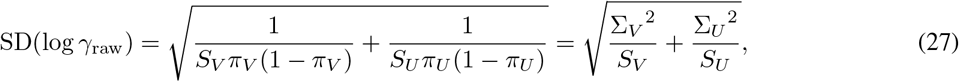

where *σ*_*V*_ = [*p*_*V*_ (1 − *p*_*V*_)]^−1*/*2^ and *σ*_*U*_ = [*p*_*U*_ (1 − *p*_*U*_)]^−1*/*2^ are the components of the true standard error and Σ_*V*_ = [*π*_*V*_ (1 − *π*_*V*_)]^−1*/*2^ and Σ_*U*_ = [*π*_*U*_ (1 − *π*_*U*_)]^−1*/*2^ are those of uncorrected standard error. We get

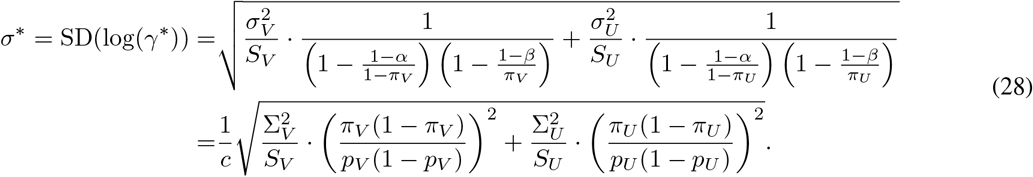

This equation indicates that the confidence intervals diverge when the true outcome is bipolarised (*p*_*V*_, *p*_*U*_ ≃ 0 or 1).

## Declarations

This research did not receive any specific grant from funding agencies in the public, commercial, or not-for-profit sectors. Authors declare that they have no competing interests.

## Footnotes

1 It has been suggested that a possible violation of this assumption occur as a result of virus interference [14], but conclusive evidence for this is currently lacking [15, 16] and the effect on VE estimates may be limited in any case [17]

2 Strictly speaking, proportion positive is a different quantity from case ratio, but it should serve as a reasonable proxy of the case ratio in most settings

## References

[1] G. De Serres, D. M. Skowronski, X. W. Wu, and C. S. Ambrose. The test-negative design: Validity, accuracy and precision of vaccine efficacy estimates compared to the gold standard of randomised placebo-controlled clinical trials. Eurosurveillance, 2013.

[2] Wakaba Fukushima and Yoshio Hirota. Basic principles of test-negative design in evaluating influenza vaccine effectiveness. Vaccine, 2017.

[3] Motoi Suzuki, Bhim Gopal Dhoubhadel, Tomoko Ishifuji, Michio Yasunami, Makito Yaegashi, Norichika Asoh, Masayuki Ishida, Sugihiro Hamaguchi, Masahiro Aoshima, Koya Ariyoshi, and Konosuke Morimoto. Serotype-specific effectiveness of 23-valent pneumococcal polysaccharide vaccine against pneumococcal pneumonia in adults aged 65 years or older: a multicentre, prospective, test-negative design study. The Lancet Infectious Diseases, 17(3):313–321, mar 2017.

[4] Rosa Prato, Francesca Fortunato, Maria Giovanna Cappelli, Maria Chironna, and Domenico Martinelli. Effective-ness of the 13-valent pneumococcal conjugate vaccine against adult pneumonia in italy: a case–control study in a 2-year prospective cohort. BMJ Open, 8(3), 2018.

[5] Kaoru Araki, Megumi Hara, Takeshi Tsugawa, Chisato Shimanoe, Yuichiro Nishida, Muneaki Matsuo, and Keitaro Tanaka. Effectiveness of monovalent and pentavalent rotavirus vaccines in japanese children. Vaccine, 36(34):5187–5193, 2018.

[6] Anna Lena Lopez, Jedas Veronica Daag, Joel Esparagoza, Joseph Bonifacio, Kimberley Fox, Batmunkh Nyambat, Umesh D Parashar, Maria Joyce Ducusin, and Jacqueline E Tate. Effectiveness of monovalent rotavirus vaccine in the Philippines. Scientific reports, 8(1):p14291, sep 2018.

[7] K Muhsen, E Anis, U Rubinstein, E Kassem, S Goren, L M Shulman, M Ephros, and D Cohen. Effectiveness of rotavirus pentavalent vaccine under a universal immunization programme in Israel, 2011–2015: a case–control study. Clinical Microbiology and Infection, 24(1):53–59, jan 2018.

[8] Evan W. Orenstein, Gaston De Serres, Michael J. Haber, David K. Shay, Carolyn B. Bridges, Paul Gargiullo, and Walter A. Orenstein. Methodologic issues regarding the use of three observational study designs to assess influenza vaccine effectiveness. International Journal of Epidemiology, 2007.

[9] Michael L. Jackson and Kenneth J. Rothman. Effects of imperfect test sensitivity and specificity on observational studies of influenza vaccine effectiveness. Vaccine, 2015.

[10] Tom De Smedt, Elizabeth Merrall, Denis Macina, Silvia Perez-Vilar, Nick Andrews, and Kaatje Bollaerts. Bias due to differential and non-differential disease- and exposure misclassification in studies of vaccine effectiveness. PLOS ONE, 2018.

[11] Sander Greenland.jBasic methods for sensitivity analysis of biases, 1996.

[12] Matthew Blackwell, James Honaker, and Gary King. A Unified Approach to Measurement Error and Missing Data: Overview and Applications. Sociological Methods and Research, 2017.

[13] M. Haber, Q. An, I. M. Foppa, D. K. Shay, J. M. Ferdinands, and W. A. Orenstein. A probability model for evaluating the bias and precision of influenza vaccine effectiveness estimates from case-control studies. Epidemiology and Infection, 143(7):1417–1426, 2015.

[14] Benjamin J. Cowling and Hiroshi Nishiura. Virus Interference and Estimates of Influenza Vaccine Effectiveness from Test-Negative Studies. Epidemiology, 2012.

[15] Increased risk of noninfluenza respiratory virus infections associated with receipt of inactivated influenza vaccine. Clinical Infectious Diseases, 2012.

[16] Maria E. Sundaram, David L. McClure, Jeffrey J. Vanwormer, Thomas C. Friedrich, Jennifer K. Meece, and Edward A. Belongia. Influenza vaccination is not associated with detection of noninfluenza respiratory viruses in seasonal studies of influenza vaccine effectiveness. Clinical Infectious Diseases, 2013.

[17] M. Suzuki, A. Camacho, and K. Ariyoshi. Potential effect of virus interference on influenza vaccine effectiveness estimates in test-negative designs. Epidemiology and Infection, 2014.

[18] V. K. Leung, B. J. Cowling, S. Feng, and Sheena G. Sullivan. Concordance of interim and final estimates of influenza vaccine effectiveness: A systematic review. 2016.

[19] Barnaby Young, Sapna Sadarangani, Lili Jiang, Annelies Wilder-Smith, and Mark I.Cheng Chen. Duration of influenza vaccine effectiveness: A systematic review, meta-analysis, and meta-regression of test-negative design case-control studies. Journal of Infectious Diseases, 2018.

[20] Effectiveness of seasonal influenza vaccine in community-dwelling elderly people: A meta-analysis of test-negative design case-control studies. The Lancet Infectious Diseases, 2014.

[21] Laurence S. Magder and James P. Hughes. Logistic regression when the outcome is measured with uncertainty. American Journal of Epidemiology, 146(2):195–203, 1997.

[22] Effectiveness of vaccine against medical consultation due to laboratory-confirmed influenza: results from a sentinel physician pilot project in British Columbia, 2004-2005. Canada Communicable Disease Report = Releve des Maladies Transmissibles au Canada, 2005.

[23] Yuki Seki, Hiroka Oonishi, Akira Onose, and Norio Sugaya. [Effectiveness of Influenza Vaccine in Adults Using A Test-negative, Case-control Design -2013/2014 and 2014/2015 Seasons-](Japanese). Kansenshogaku zasshi. The Journal of the Japanese Association for Infectious Diseases, 2016.

[24] Yuki Seki, Akira Onose, and Norio Sugaya. Influenza vaccine effectiveness in adults based on the rapid influenza diagnostic test results, during the 2015/16 season. Journal of Infection and Chemotherapy, 2017.

[25] Nobuo Saito, Kazuhiro Komori, Motoi Suzuki, Kounosuke Morimoto, Takayuki Kishikawa, Takahiro Yasaka, and Koya Ariyoshi. Negative impact of prior influenza vaccination on current influenza vaccination among people infected and not infected in prior season: A test-negative case-control study in Japan. Vaccine, 2017.

[26] Masayoshi Shinjoh, Norio Sugaya, Yoshio Yamaguchi, Noriko Iibuchi, Isamu Kamimaki, Anna Goto, Hisato Kobayashi, Yasuaki Kobayashi, Meiwa Shibata, Satoshi Tamaoka, Yuji Nakata, Atsushi Narabayashi, Mitsuhiro Nishida, Yasuhiro Hirano, Takeshi Munenaga, Kumiko Morita, Keiko Mitamura, and Takao Takahashi. Inactivated influenza vaccine effectiveness and an analysis of repeated vaccination for children during the 2016/17 season. Vaccine, 2018.

[27] Soichiro Ando. Effectiveness of quadrivalent influenza vaccine based on the test-negative control study in children during the 2016–2017 season. Journal of Infection and Chemotherapy, 2018.

[28] Norio Sugaya, Masayoshi Shinjoh, Yuji Nakata, Kenichiro Tsunematsu, Yoshio Yamaguchi, Osamu Komiyama, Hiroki Takahashi, Keiko Mitamura, Atsushi Narabayashi, and Takao Takahashi. Three-season effectiveness of inactivated influenza vaccine in preventing influenza illness and hospitalization in children in Japan, 2013-2016. Vaccine, 2018.

[29] Sheena G. Sullivan, Shuo Feng, and Benjamin J. Cowling. Potential of the test-negative design for measuring influenza vaccine effectiveness: A systematic review. Expert Review of Vaccines, 2014.

[30] Comparison of two control groups for estimation of oral cholera vaccine effectiveness using a case-control study design. Vaccine, 2017.

[31] Joseph A. Lewnard, Christine Tedijanto, Benjamin J. Cowling, and Marc Lipsitch. Measurement of vaccine direct effects under the test-negative design. American Journal of Epidemiology, 2018.

[32] Direk Limmathurotsakul, Elizabeth L. Turner, Vanaporn Wuthiekanun, Janjira Thaipadungpanit, Yupin Suputtamongkol, Wirongrong Chierakul, Lee D. Smythe, Nicholas P.J. Day, Ben Cooper, and Sharon J. Peacock. Fool’s gold: Why imperfect reference tests are undermining the evaluation of novel diagnostics: A reevaluation of 5 diagnostic tests for leptospirosis. Clinical Infectious Diseases, 2012.

[33] Christiana A. Naaktgeboren, Loes C.M. Bertens, Maarten van Smeden, Joris A.H. de Groot, Karel G.M. Moons, and Johannes B. Reitsma. Value of composite reference standards in diagnostic research. BMJ (Clinical research ed.), 2013.

[34] Suzan R. Kahn, Elham Rahme, Lisa M. Lix, Mark Burman, Geneviève Lefebvre, Jiayi Ni, Kaberi Dasgupta, Yves Laflamme, Greg Berry, Ronald Dimentberg, Denis Talbot, and Alain Cirkovic. Comparing external and internal validation methods in correcting outcome misclassification bias in logistic regression: A simulation study and application to the case of postsurgical venous thromboembolism following total hip and knee arthroplasty. Pharmacoepidemiology and Drug Safety, 2018.

